# The wellbeing paradox: High resilience and psychological distress in the transition out of UK policing

**DOI:** 10.64898/2026.03.14.26348403

**Authors:** Eleftheria Vaportzis, Warren Edwards

## Abstract

This study investigated the wellbeing of UK police officers transitioning out of service, examining retirees, early leavers, and those within 12 months of retirement (N = 370). Using the Job Demands–Resources model, the research identifies a ‘wellbeing paradox’: leavers demonstrate high resilience and subjective wellbeing alongside significantly elevated psychological distress compared to general population norms. Findings reveal that recently retired (≤5 years) and soon-to-retire groups are particularly vulnerable, reporting lower quality of life and higher distress than long-term retirees. Perceived organisational support and resilience emerged as critical buffers against the psychological burden of a policing career. However, participants identified significant unmet needs for career, financial, and mental health guidance during the transition. The study highlights that the anticipatory retirement period is an acute window of vulnerability, suggesting that proactive, targeted organisational interventions are essential to mitigate the lasting psychological burden of policing and ensure successful civilian transitions.

---

Policing is a stressful profession, characterised by repeated exposure to traumatic events, heavy workloads, and organisational pressures that can negatively impact wellbeing over the course of a career (Oliver, Thomas, Neil, Moll, & Copeland, 2023). Police officers often spend their career in environments that demand emotional control, resilience, and commitment to public service (Hopkins, Dowell, & Flitton, 2023). Leaving policing, whether through retirement, resignation, or ill health, often presents substantial challenges for officers (Jones, Hesketh, & Noble, 2025b). These transitions can involve lingering psychological effects of trauma (Porter & Lee, 2024), financial pressures linked to organisational reforms and pension uncertainty (Jones et al., 2025b), occupational shifts as officers move away from structured roles (Demou, Hale, & Hunt, 2020), loss of peer networks, alongside identity disruption (Charman & Tyson, 2023), and cultural challenges such as stigma and perceptions of organisational fairness (Phythian et al., 2022). Leaving policing is not a single event but a multidimensional process, influenced by organisational culture, role identity, and individual circumstances, with markedly different experiences across cohorts (Charman & Tyson, 2024; Tyson & Charman, 2025).

The past decade has seen significant transformation in UK policing. Workforce austerity, rising demand, organisational reform, and heightened scrutiny have intensified job demands while constraining resources (Topping, 2022). Pension reforms, including the 2015 Police Pension Scheme and subsequent legal adjustments, have complicated retirement trajectories, generating uncertainty, and extending working lives (Home Office, 2019; NARPO, 2019). Critical to this uncertainty is the ‘McCloud remedy’, the legal rectification of age discrimination within the 2015 reforms (Lord Chancellor v McCloud and Others, 2018). The ‘remedy’ is a legal requirement for the government to remove age discrimination by allowing affected members to choose between their legacy and reformed pension schemes for the remedy period. Its implementation has triggered significant administrative chaos, characterised by delayed Remediable Service Statements and complex retrospective tax liabilities (House of Commons Library, 2024). This has transformed a technical correction into a source of profound psychological and financial distress, leaving many officers in a state of limbo regarding their retirement dates and future security (Jones et al., 2025b). Traditionally, pensions provided stability by supporting retention and easing transition; however, recent reforms have diminished this role, creating uncertainty and affecting wellbeing both during service and after exit (Jones et al., 2025b; Metfriendly, 2025). In response to these organisational changes, national initiatives including the Blue Light Wellbeing Framework and the National Police Wellbeing Service, demonstrate a growing recognition of the psychological costs of policing (Oscar Kilo, 2024a, 2024b). Yet evidence suggests that implementation remains uneven, constrained by cultural stigma and concerns over organisational justice, leaving many officers with unmet needs (Porter & Lee, 2024; Lennie et al., 2025).

Patterns of police exit increasingly reflect the cumulative impact of these organisational and cultural factors. While voluntary resignation has attracted attention, similar dynamics such as emotional exhaustion, lack of voice, organisational injustice, and erosion of commitment, also shape retirement and medical□health exits (Charman & Bennett, 2022; Lennie et al., 2025). Officers frequently express a strong sense of vocational identity and commitment to the policing role, yet this often coexists with growing dissatisfaction toward organisational practices and leadership (Hoggett, Redford, Toher, & White, 2019). This tension can disrupt identity, diminish wellbeing, and make it harder to rebuild a sense of purpose after leaving the Force (Roach, Curran, Cartwright, & McCarthy, 2025). This phenomenon may contribute to a ‘wellbeing paradox’ wherein former officers maintain high functional wellbeing despite carrying a significant and lasting psychological burden from their career (Syed et al., 2020).

The Job Demands–Resources (JD-R) model (Bakker & Demerouti, 2007) provides a useful framework for understanding these processes: chronic demands such as trauma exposure, workload, and emotional labour predict strain, while resources such as organisational support, autonomy, and resilience buffer these effects (Hesketh & Tehrani, 2019). Extending the JD-R model into exit contexts highlights how the legacy of demands and the availability (or absence) of resources continue to impact wellbeing after service ends.

In this context, examining the wellbeing of police leavers provides critical insight into how officers experience the transition out of service across different stages. By considering those preparing to leave, as well as those who have already exited, the study captures both anticipatory concerns and lived experiences. This approach also enables exploration of cohort effects, particularly whether officers who retired under recent conditions, such as pension reforms, differ from earlier cohorts who left under more stable institutional arrangements. The study’s primary question is: How do retired officers who left in the past five years differ from those who retired six or more years ago in terms of wellbeing, resilience, and perceived organisational support? Secondary questions examine whether differences exist between current officers preparing to leave, retirees, and those who left the Force before retirement. By addressing these questions, the study aims to generate evidence that can inform police organisations, representative bodies, and policymakers in developing targeted, evidence□based interventions to support officers before, during, and after exit, ensuring wellbeing is maintained throughout the transition to civilian life.

## Method

### Recruitment

A cross-sectional survey was designed, following guidelines by Kelley et al. (2003). Eligibility criteria included being a former police officer in the UK, either retired or having left before retirement, or planning to retire within the next 12 months. The survey was distributed online via Online Surveys (www.onlinesurveys.ac.uk) between November 2025 and January 2026. Data collection continued until two consecutive days passed without any new completions. Duplicate entries were minimised using IP tracking and timestamp checks provided by the Online Surveys platform. Participants were recruited using multiple outreach methods targeting police officers. Recruitment materials were disseminated via social media posts (e.g., Facebook, X, Instagram) and community groups related to police officers in the UK. The study was promoted through newsletters, websites, and mailing lists of relevant organisations across the UK [e.g., National Association of Retired Police Officers (NARPO), Police Superintendents’ Association, College of Policing]. These organisations were contacted directly and received study materials including a poster with the survey link for dissemination. Snowball sampling was encouraged, and participants were invited to share the survey with others who may be eligible. All recruitment materials included eligibility criteria, ethics approval reference, and contact details for the research team. Ethics approval was granted by the Humanities, Social, and Health Sciences Research Ethics Panel at the University of Bradford (Reference Number E1350). All participants provided written informed consent.

### Survey design

#### Demographic and service questions

Demographic questions included participants’ age, sex assigned at birth, ethnic group, highest education completed, marital status, and current living situation. Service questions included UK service region, current status in relation to police service, highest rank attained, years of service, voluntariness of exit, and retirement length. Year of service and retirement length were converted from months to decimal years (e.g., 1 month = 0.08) to ensure linear consistency in the models.

#### Short Warwick-Edinburgh Mental Well-Being Scale (SWEMWBS; Stewart-Brown et al., 2009)

SWEMWBS is a validated 7-item measure of subjective wellbeing. Items are positively worded statements (e.g., “I’ve been feeling optimistic about the future”) rated on a 5-point Likert scale (1 = None of the time – 5 = All of the time). Scores range between 7 and 35, with higher scores indicating better subjective wellbeing. Cronbach’s α in the current study was excellent (.92).

#### EUROHIS-QOL 8-Item Index (Schmidt, Mühlan, & Power, 2006)

EUROHIS-QOL 8-Item Index is a cross-culturally validated measure of quality of life derived from the WHOQOL questionnaire. It comprises 8 items covering physical, psychological, social, and environmental domains rated on a 5-point Likert scale. Scores range between 8 and 40, with higher scores indicating better quality of life. Cronbach’s α was excellent (.91).

#### Depression, Anxiety, and Stress Scale – 21 items (DASS-21; Lovibond, 1995)

DASS-21 is a validated measure that assesses feelings of depression, anxiety, and stress. It comprises three 7-item scales. Participants are asked to indicate whether each statement has applied to them during the last week. Responses are rated on a 4-point Likert-type scale (0□=□Did not apply to me at all - 3□=□Applied to me a lot/most of the time). Scores are summed and multiplied by two. The total score for each scale ranges between 0 and 42. Higher scores indicate a higher level of the respective construct. The total DASS-21 score is the sum of the three scales and can range between 0 and 126 with higher scores indicating a higher level of general psychological distress. Cronbach’s α was excellent for the overall scale (.96), Depression (.95), and Stress (.92), and good for Anxiety (.87).

#### Brief Resilience Scale (BRS; Smith et al., 2008)

BRS is a 6-item measure designed to assess the ability to recover from stress. Items (e.g., *“I tend to bounce back quickly after hard times”*) are rated on a 5-point Likert scale (1 = Strongly disagree – 5 = Strongly agree). To ensure that higher scores consistently reflected a greater capacity for resilience, the three negatively phrased items were reverse-coded. Scores range between 6 and 30, with higher scores indicating greater resilience. Cronbach’s α was acceptable (.71).

#### Adapted Perceived Organizational Support (Barnes, Nickerson, Adler, & Litz, 2013)

This adapted version of the Perceived Organizational Support scale modifies 6 items to reflect military contexts (e.g., *“My force cares/cared about my wellbeing”*). It assesses the extent to which service members feel valued and supported by their organisation. Items are rated on a Likert scale (1 = Strongly disagree – 5 = Strongly agree). Scores range between 6 and 42 with higher scores indicating stronger perceived organisational support. Cronbach’s α was excellent (.93).

#### Trauma exposure

To capture the presence of duty-related traumatic exposure, participants were asked a single screening question adapted from trauma exposure assessments (Blevins, Weathers, Davis, Witte, & Domino, 2015): “Have you experienced a distressing or traumatic event in the course of your police duties (e.g., serious accident, use of force, violent death, or threat to your life) that has continued to affect you emotionally?” (1 = Yes, 2 = No, 3 = Don’t know, 4 = Prefer not to say). This item was used to capture the presence of duty□related traumatic exposure rather than to provide a full symptom severity score.

#### Shift schedule satisfaction

Participants responded to a study□specific item designed to capture satisfaction with shift schedule: “How satisfied were you with your shift schedule during your last 3 years of service? (If currently serving, please reflect on your current schedule; 1 = Very dissatisfied – 5 = Very satisfied).

#### Additional support needs

Participants were asked to respond to three study□specific items designed to capture perceived support needs during and after police service. Each item was rated on a 5□point Likert scale (1 = Strongly disagree – 5 = Strongly agree). The items were: “I would have welcomed career/second career advice”, “I would have welcomed effective signposting to welfare and mental health services”, and “I would have welcomed pension/financial advice”. Responses provided an indication of the extent to which participants felt that additional guidance and resources would have been beneficial. These items were developed for the purposes of this study to explore unmet support needs. Higher scores reflect stronger agreement with the statement and therefore greater perceived need for the specified type of support.

### Data analysis

Statistical analyses were performed in SPSS v. 28 (IBM, 2021) with alpha set at .05. Potential outliers were identified using Z-score cut-offs (± 3.29; Tabachnick & Fidell, 2013). While two significant outliers were identified on the DASS-21 Anxiety scale, the scores were deemed plausible, did not influence the overall results, and, therefore, they were retained. Bivariate relationships were assessed using Pearson’s correlation coefficients.

To examine group differences in quality of life, subjective wellbeing, and psychological distress among retired officers, early leavers, and soon-to-retire officers, a Multivariate Analysis of Variance (MANOVA) was conducted. Due to violations of the homogeneity of variance-covariance matrices (assessed using Box’s M test), and the inherent imbalance in group sizes, Welch’s one-way ANOVA with Games-Howell post-hoc tests were employed for all continuous pairwise comparisons. These methods were chosen as they do not assume equal variances or group sizes. To ensure further robustness, bootstrapped estimates (2,000 samples, 95% CI) were calculated for all MANOVAs and subsequent regressions to account for non-normal score distributions.

Categorical variables, such as rank and support needs, were evaluated using Chi-square tests for independence. Likelihood ratios were reported where expected cell counts were low (< 5). Post-hoc analysis for categorical data was performed using adjusted standardised residuals. For the retired group specifically, long-term adaptation was evaluated using Mann-Whitney U tests to compare those retired for five years or less with those retired for six or more years. To test the JD-R model, a hierarchical multiple regression was conducted with subjective wellbeing, quality of life, and psychological distress as outcomes. Predictors were entered in three steps: (1) contextual factors, including transition status (i.e., retired, early leaver, soon-to-retire); (2) job demands, including trauma exposure and shift satisfaction; and (3) resources, including perceived organisational support and resilience. This structure allows for the examination of the buffering hypothesis, determining if ‘Resources’ mitigate the relationship between chronic occupational ‘Demands’ and post-service outcomes. Models were checked for outliers (standardised residuals ± 3, Cook’s distance >1) and multicollinearity (Tolerance <.01, VIF >10).

The survey employed a forced-response design, resulting in no missing data for validated scales. A priori power analysis using G*Power (Faul, Erdfelder, Buchner, & Lang, 2009) indicated that a total sample of 647 participants would be required to detect a small effect (f^2^ = 0.02) with α = .05 and power = .80. Given the final sample of N = 370, the study is sufficiently powered to detect medium-to-large effects (f^2^ = 0.15), while comparisons involving the smaller ‘early leaver’ (n = 42) and ‘soon-to-retire’ (n = 39) groups should be treated as exploratory.

## Results

### Sample demographics

Participants included retired officers (n = 289, 78.1%), comprising those who retired on pensionable years, took early retirement, or retired on medical grounds; early leavers (n = 42, 11.4%), comprising those who left the Force prior to reaching retirement age; and soon-to-retire officers (n = 39, 10.5%), comprising active-duty officers planning to leave the Force within the following 12 months. The total sample comprised 271 males (73.2%), 98 females (26.5%), and 1 participant who preferred not to report their sex (0.3%). The mean age of the sample was 61.92 years (*SD* = 10.48, range 24-93). The majority of respondents identified as ‘White English, Welsh, Scottish, Northern Irish or Irish’ (n = 355, 95.9%), and served in England or Wales (n = 341, 92.2%), with the remaining participants representing Scotland (n = 16, 4.3%) and Northern Ireland (n = 13, 3.5%). Among the participants who had already retired, nearly 30% (n = 90, 29.3%) had done so within the last 5 years, with the remaining having retired for 6 or more years (n = 217, 70.7%). Most participants were married or in a civil partnership (n = 287, 77.6%), while others were divorced (n = 32, 8.6%), single (n = 19, 5.1%), or widowed (n = 12, 3.2%). The majority of the sample lived with a partner or spouse (n = 244, 65.9%) or with a partner/spouse and children (n = 63, 17.0%), while 11.6% (n = 43) reported living alone. Educational attainment varied, with the largest groups having completed secondary school (n = 91, 24.6%), a vocational or technical qualification (n = 71, 19.2%), college (n = 70, 18.9%), or a Bachelor’s degree (n = 68, 18.4%). Finally, for those who had already left the Force, the majority indicated their departure was voluntary (n = 291, 78.6%), compared to 8.4% (n = 31) whose departure was involuntary. Further service characteristics are compared by retirement group in Table 1.

**Table 1.**
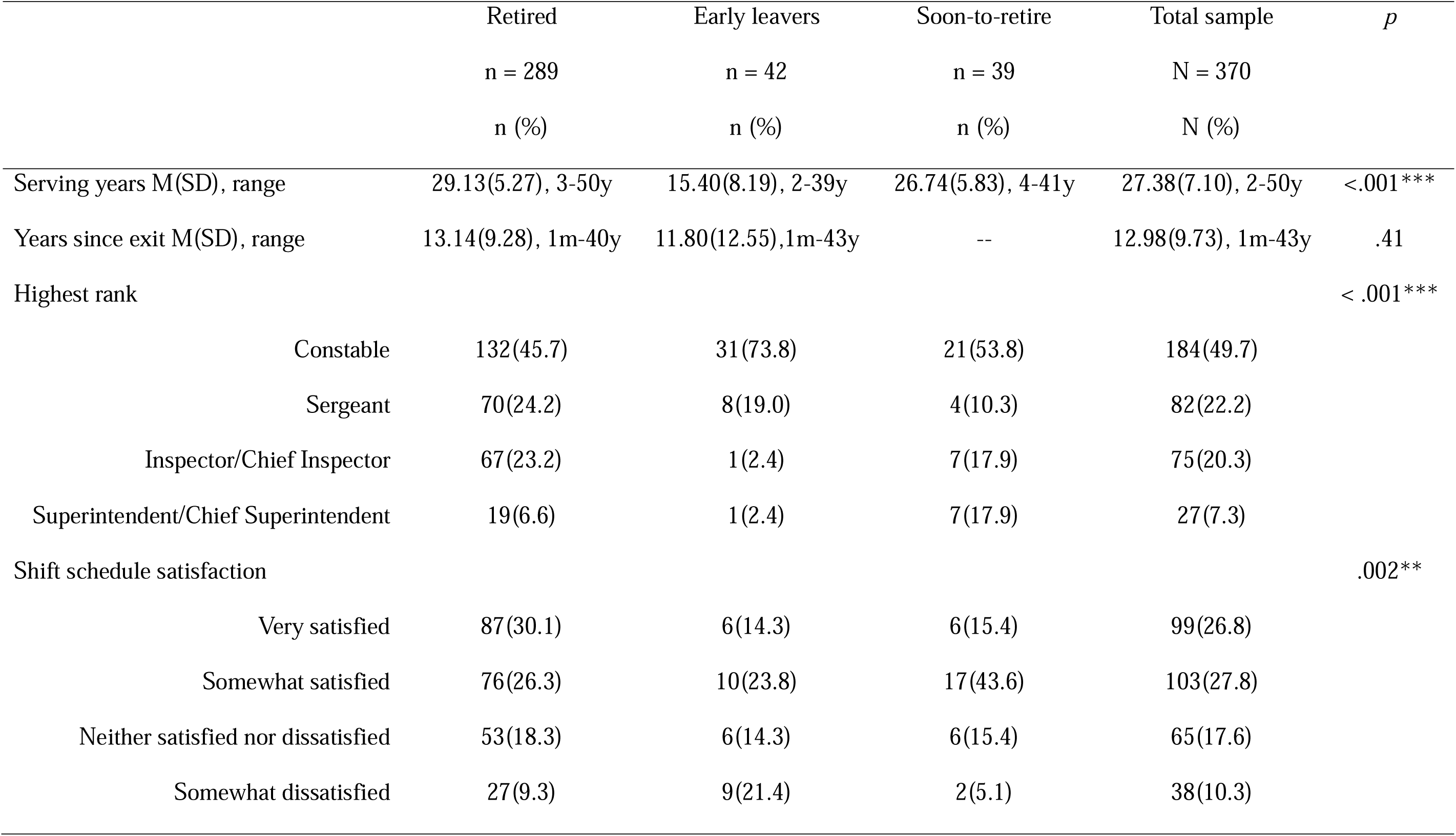
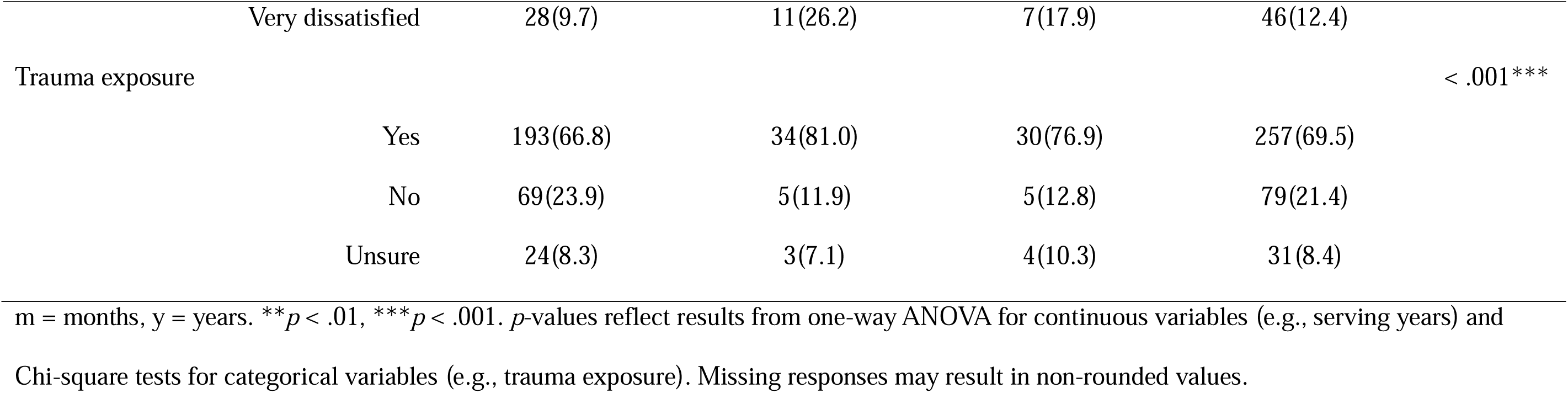
Service characteristics and occupational impact by group.

### Differences in wellbeing, psychological distress, and resources by time since leaving policing

Mann-Whitney U tests revealed that participants who retired for five years or less reported significantly lower scores than those retired for six years or more across subjective wellbeing (U = 11888, *p* = .01), quality of life (U = 12602, *p* < .001), and perceived organisational support (U = 12281, *p* < .001). In addition, the more recently retired group reported significantly higher levels of depression (U = 21215, *p* = .002), anxiety (U = 31013, *p* = .001), and stress (U = 7372, *p* = .001). No significant differences were observed between the two groups regarding resilience (U = 9163, *p* = .38).

### Differences in wellbeing, psychological distress, and resources by exit pathway

MANOVA showed significant differences between the groups across all dependent variables (*p* < .001). The largest effect was observed for quality of life, *F*(2, 367) = 17.01, *p* < .001, η² = .08, followed by subjective wellbeing, *F*(2, 367) = 13.80, *p* < .001, η² = .06. Significant group effects were also found for stress, *F*(2, 367) = 11.74, *p* < .001, η² = .05, depression, *F*(2, 367) = 11.12, *p* < .001, η² = .05, and anxiety, *F*(2, 367) = 9.89, *p* < .001, η² = .05. Organisational support also differed significantly between groups, *F*(2, 367) = 6.38, *p* = .002, η² = .03, as well as resilience, *F*(2, 367) = 3.84, *p* = .02, η² = .02.

Post-hoc tests showed that soon-to-retire participants had significantly lower scores than those who were retired on quality of life (*p* < .001) and wellbeing (*p* < .001). This group also reported significantly higher stress (*p* = .001) and depression scores (*p* = .01) than retirees. Early leavers had significantly lower quality of life scores (*p* = .002) and significantly higher depression (*p* = .041), anxiety (*p* = .022), and stress scores (*p* = .014) than those who were retired. For organisational support, early leavers reported significantly lower scores than retired officers (*p* < .001), while no significant differences were found between soon□to□retire participants and either of the other groups. No significant differences were found between those soon-to-retire and those who left the Force before retirement across most of the wellbeing or psychological distress measures (all *p* > .05), with the exception of wellbeing, where soon-to-retire participants scored significantly lower than early leavers (*p* = .03). Resilience did not differ significantly between any of the three groups (all *p* > .05).

In comparison with normative data, the whole sample showed significantly poorer mental health across all psychological distress measures. Depression, *t*(368) = 8.00, *p* < .001, anxiety, *t*(368) = 7.91, *p* < .001, and stress, *t*(368) = 5.54, *p* < .001, were all elevated relative to population norms (Crawford & Henry, 2003). Conversely, the sample showed better scores than normative values in both subjective wellbeing, *t*(368) = 3.76, *p* < .001 (Ng Fat, Scholes, Boniface, Mindell, & Stewart-Brown, 2017), and resilience, *t*(368) = 126.35, *p* < .001 (Smith et al., 2008). No significant difference was found for quality of life, *t*(368) = –0.01, *p* = .99 (Schmidt et al., 2006).

### Predicting post-service adjustment: A JD-R perspective

Hierarchical multiple regression was employed to determine the extent to which contextual factors and JD-R variables predicted post-service adjustment. A consistent pattern emerged across wellbeing, quality of life, and the three measures of psychological distress (see Tables 2-6). In all models, perceived organisational support emerged as a significant predictor across all outcomes, including improved wellbeing (β = .27, *p* < .001) and reduced stress (β = -.25, *p* < .001). Once demands and resources were included, the effect of retirement length consistently remained negligible and non-significant across all measures (*p* > .05).

**Table 2.**
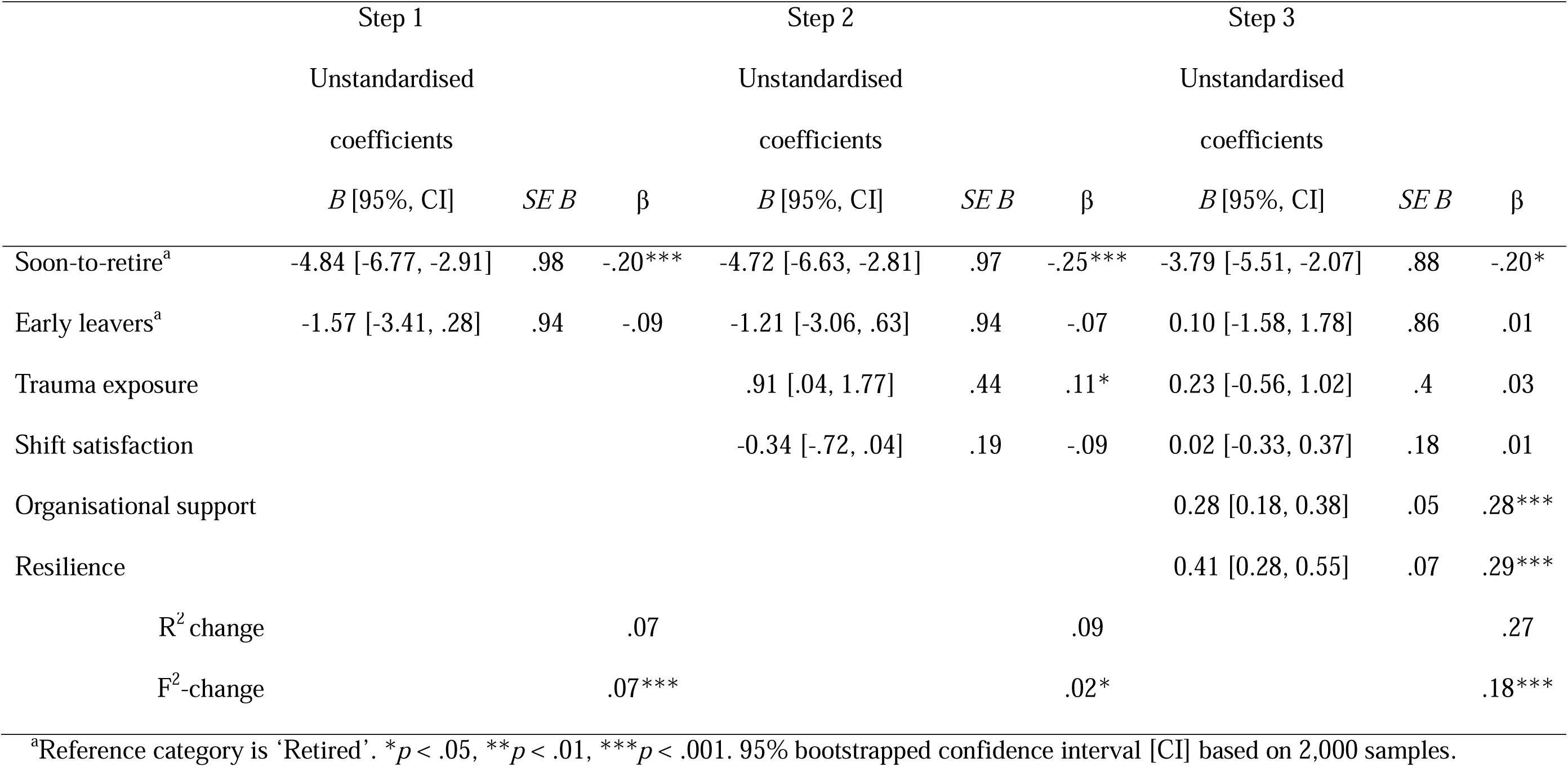
Hierarchical regression analysis for wellbeing.

|  | Step 1 |  |  | Step 2 |  |  | Step 3 |  |  |
| --- | --- | --- | --- | --- | --- | --- | --- | --- | --- |
|  | Unstandardised |  |  | Unstandardised |  |  | Unstandardised |  |  |
|  | coefficients |  |  | coefficients |  |  | coefficients |  |  |
| | *B* [95%, CI] | *SE B* | $\beta$ | *B* [95%, CI] | *SE B* | $\beta$ | *B* [95%, CI] | *SE B* | $\beta$ |
| Soon-to-retire<sup>a</sup> | -4.84 [-6.77, -2.91] | .98 | -.20*** | -4.72 [-6.63, -2.81] | .97 | -.25*** | -3.79 [-5.51, -2.07] | .88 | -.20* |
| Early leavers<sup>a</sup> | -1.57 [-3.41, .28] | .94 | -.09 | -1.21 [-3.06, .63] | .94 | -.07 | 0.10 [-1.58, 1.78] | .86 | .01 |
| Trauma exposure |  |  |  | .91 [.04, 1.77] | .44 | .11* | 0.23 [-0.56, 1.02] | .4 | .03 |
| Shift satisfaction |  |  |  | -0.34 [-.72, .04] | .19 | -.09 | 0.02 [-0.33, 0.37] | .18 | .01 |
| Organisational support |  |  |  |  |  |  | 0.28 [0.18, 0.38] | .05 | .28*** |
| Resilience |  |  |  |  |  |  | 0.41 [0.28, 0.55] | .07 | .29*** |
| R<sup>2</sup> change |  |  | .07 |  |  | .09 |  |  | .27 |
| F<sup>2</sup>-change |  |  | .07*** |  |  | .02* |  |  | .18*** |

<sup>a</sup>Reference category is 'Retired'. \* $p < .05$ , \*\* $p < .01$ , \*\*\* $p < .001$ . 95% bootstrapped confidence interval [CI] based on 2,000 samples.

**Table 3.** Hierarchical regression analysis for quality of life.

|  | Step 1 |  |  | Step 2 |  |  | Step 3 |  |  |
| --- | --- | --- | --- | --- | --- | --- | --- | --- | --- |
|  | Unstandardised |  |  | Unstandardised |  |  | Unstandardised |  |  |
|  | coefficients |  |  | coefficients |  |  | coefficients |  |  |
| | *B* [95%, CI] | *SE B* | $\beta$ | *B* [95%, CI] | *SE B* | $\beta$ | *B* [95%, CI] | *SE B* | $\beta$ |
| Soon-to-retire<sup>a</sup> | -5.35 [-7.61, -3.09] | 1.15 | -.23*** | -5.17 [-7.40, -2.94] | 1.14 | -.23*** | -4.14 [-6.17, -2.10] | 1.04 | -.18*** |
| Early leavers<sup>a</sup> | -4.27 [-6.43, -2.10] | 1.10 | -.20*** | -3.75 [-5.90, -1.60] | 1.10 | -.17*** | -2.40 [-4.38, -.41] | 1.01 | -.11** |
| Trauma exposure |  |  |  | 1.35 [.35, 2.36] | .51 | .13** | .63 [-.30, 1.56] | .48 | .06 |
| Shift satisfaction |  |  |  | -.47 [-.91, -.03] | .22 | -.11* | -.09 [-.50, .32] | .21 | -.02 |
| Organisational support |  |  |  |  |  |  | .26 [.15, .38] | .06 | .22*** |
| Resilience |  |  |  |  |  |  | .51 [.35, .67] | .08 | .30*** |
| R<sup>2</sup> change |  |  | .08 |  |  | .11 |  |  | .27 |
| F<sup>2</sup>-change |  |  | .08*** |  |  | .03*** |  |  | .16*** |

<sup>a</sup>Reference category is 'Retired'. \* $p < .05$ , \*\* $p < .01$ , \*\*\* $p < .001$ . 95% bootstrapped confidence interval [CI] based on 2,000 samples.

**Table 4.** Hierarchical regression analysis for depression.

|  | Step 1 |  |  | Step 2 |  |  | Step 3 |  |  |
| --- | --- | --- | --- | --- | --- | --- | --- | --- | --- |
|  | Unstandardised |  |  | Unstandardised |  |  | Unstandardised |  |  |
|  | coefficients |  |  | coefficients |  |  | coefficients |  |  |
| | *B* [95%, CI] | *SE B* | $\beta$ | *B* [95%, CI] | *SE B* | $\beta$ | *B* [95%, CI] | *SE B* | $\beta$ |
| Soon-to-retire<sup>a</sup> | 6.90 [2.11, 11.35] | 2.33 | .19** | 6.61 [1.97, 10.80] | 2.25 | .18* | 4.87 [.70, 8.81] | 2.06 | .13* |
| Early leavers<sup>a</sup> | 5.31 [1.34, 9.66] | 2.11 | .15* | 4.48 [.32, 8.99] | 2.17 | .13** | 2.09 [-1.62, 6.02] | 1.94 | .06 |
| Trauma exposure |  |  |  | -2.20 [-4.01, -.42] | .91 | -.13** | -.95 [-2.59, .68] | .84 | -.06 |
| Shift satisfaction |  |  |  | .75 [.04, 1.52] | .37 | .1* | .09 [-.55, .75] | .33 | .01 |
| Organisational support |  |  |  |  |  |  | -.49 [-.68, -.29] | .1 | -.26*** |
| Resilience |  |  |  |  |  |  | -.80 [-1.07, -.55] | .13 | -.3*** |
| R<sup>2</sup> change |  |  | .05 |  |  | .08 |  |  | .26 |
| F<sup>2</sup>-change |  |  | .05*** |  |  | .03** |  |  | .18*** |

<sup>a</sup>Reference category is 'Retired'. \* $p < .05$ , \*\* $p < .01$ , \*\*\* $p < .001$ . 95% bootstrapped confidence interval [CI] based on 2,000 samples.

**Table 5.** Hierarchical regression analysis for anxiety.

|  | Step 1 |  |  | Step 2 |  |  | Step 3 |  |  |
| --- | --- | --- | --- | --- | --- | --- | --- | --- | --- |
|  | Unstandardised |  |  | Unstandardised |  |  | Unstandardised |  |  |
|  | coefficients |  |  | coefficients |  |  | coefficients |  |  |
| | *B* [95%, CI] | *SE B* | $\beta$ | *B* [95%, CI] | *SE B* | $\beta$ | *B* [95%, CI] | *SE B* | $\beta$ |
| Soon-to-retire<sup>a</sup> | 4.20 [1.29, 7.10] | 1.48 | .15** | 3.99 [1.12, 6.86] | 1.46 | .14** | 2.79 [.11, 5.46] | 1.37 | .10* |
| Early leavers<sup>a</sup> | 4.97 [2.19, 7.75] | 1.42 | .18* | 4.36 [1.59, 7.13] | 1.41 | .16** | 2.74 [.13, 5.35] | 1.33 | .10** |
| Trauma exposure |  |  |  | -1.41 [-2.71, -.11] | .66 | -.11* | -.56 [-1.78, .67] | .62 | -.04 |
| Shift satisfaction |  |  |  | .62 [.06, 1.19] | .29 | .11* | .17 [-.37, .71] | .28 | .03 |
| Organisational support |  |  |  |  |  |  | -.33 [-.48, -.17] | .08 | -.22*** |
| Resilience |  |  |  |  |  |  | -.57 [-.78, -.37] | .11 | -.27*** |
| R<sup>2</sup> change |  |  | 0.05 |  |  | .08 |  |  | .21 |
| F<sup>2</sup>-change |  |  | .05*** |  |  | .03** |  |  | .14*** |

<sup>a</sup>Reference category is 'Retired'. \* $p < .05$ , \*\* $p < .01$ , \*\*\* $p < .001$ . 95% bootstrapped confidence interval [CI] based on 2,000 samples.

**Table 6.** Hierarchical regression analysis for stress.

|  | Step 1 |  |  | Step 2 |  |  | Step 3 |  |  |
| --- | --- | --- | --- | --- | --- | --- | --- | --- | --- |
|  | Unstandardised |  |  | Unstandardised |  |  | Unstandardised |  |  |
|  | coefficients |  |  | coefficients |  |  | coefficients |  |  |
| | *B* [95%, CI] | *SE B* | $\beta$ | *B* [95%, CI] | *SE B* | $\beta$ | *B* [95%, CI] | *SE B* | $\beta$ |
| Soon-to-retire<sup>a</sup> | 6.71 [3.20, 10.21] | 1.79 | .13*** | 6.35 [2.93, 9.77] | 1.75 | .18*** | 4.63 [1.56, 7.69] | 1.56 | .13*** |
| Early leavers<sup>a</sup> | 5.40 [2.04, 8.75] | 1.71 | .16** | 4.51 [1.21, 7.81] | 1.68 | .14** | 2.20 [-.79, 5.19] | 1.53 | .07** |
| Trauma exposure |  |  |  | -3.14 [-4.69, -1.59] | .79 | -.20*** | -1.93 [-3.33, -.53] | 0.72 | -.12 |
| Shift satisfaction |  |  |  | .57 [-.10, 1.25] | .35 | .08 | -.07 [-.68, .55] | .32 | -.01** |
| Organisational support |  |  |  |  |  |  | -.46 [-.64, -.29] | .09 | -.25*** |
| Resilience |  |  |  |  |  |  | -.82 [-1.05, -.58] | .12 | -.32*** |
| R<sup>2</sup> change |  |  | .06 |  |  | .11 |  |  | .30 |
| F<sup>2</sup>-change |  |  | .06*** |  |  | .05*** |  |  | .19*** |

<sup>a</sup>Reference category is 'Retired'. \* $p < .05$ , \*\* $p < .01$ , \*\*\* $p < .001$ . 95% bootstrapped confidence interval [CI] based on 2,000 samples.

Being in the soon-to-retire group remained a significant predictor of poorer outcomes in most final models, regardless of other resources. This group demonstrated significantly lower wellbeing (β = -.11, *p* = .003) and quality of life (β = -.14, *p* < .001) compared to the retired group. Additionally, resilience was consistently associated with better outcomes; higher resilience scores significantly predicted higher wellbeing (β = .15, *p* < .001) and quality of life (β = .19, *p* < .001), as well as lower levels of depression (β = .19, *p* < .001), anxiety (β = -.21, *p* < .001), and stress (β = -.23, *p* < .001).

To further explore the practical implications of these findings, participants’ perceived support needs were examined. Descriptive analysis revealed that the majority of the sample would have welcomed additional assistance in the form of career advice (61.1%), financial or pension advice (57.3%), and effective signposting to welfare and mental health services (52.7%). Correlation analyses further demonstrated that these unmet support needs were significantly associated with poorer adjustment outcomes across the sample. Specifically, a stronger desire for welfare signposting showed the most robust relationships with negative outcomes, correlating significantly with higher levels of depression (r = .45, *p* < .001), anxiety (r = .39, *p* < .001), and stress (r = .51, *p* < .001), as well as lower subjective wellbeing (r = -.40, *p* < .001) and quality of life (r = -.46, *p* < .001). Significant correlations were also observed for the desire for career and financial advice across all measures of wellbeing and distress (all *p* < .01).

One-way ANOVAs were conducted to determine if these needs differed significantly across the three groups. Results indicated significant group differences for all three items: career advice, *F*(2, 367) = 13.51, *p* < .001; welfare signposting, *F*(2, 367) = 13.86, *p* < .001; and financial advice, *F*(2, 365) = 10.96, *p* < .001. Post-hoc comparisons showed that the soon-to-retire group reported a significantly higher need for career advice (p < .001), and financial and pension advice (p < .001) compared to those already retired. Furthermore, early leavers expressed a significantly higher need for welfare and mental health signposting than both the retired (p < .001) and soon-to-retire (p = .04) groups. Early leavers also reported a significantly higher desire for career advice compared to retired officers (p = .004).

## Discussion

This study investigated the wellbeing, quality of life, and psychological distress of UK police leavers and those approaching retirement. By examining different exit pathways, the research highlights the significant impact of organisational factors and timing on the transition to civilian life. Our primary question investigated differences between retired officers who left the Force in the past five years and those who retired six or more years ago in terms of wellbeing, resilience, and perceived organisational support. The analysis revealed significant differences between cohorts based on retirement timing. Officers who retired within the last five years reported significantly lower subjective wellbeing, quality of life, and perceived organisational support than those who retired six or more years ago. They also reported significantly higher levels of depression, anxiety, and stress. These findings support the notion that recent organisational shifts, such as austerity and pension reforms, have created a more challenging transition environment for officers navigating the modern landscape.

The transition status (i.e., retired, soon-to-retire, or early leaver) significantly influenced outcomes. Soon-to-retire participants appeared most vulnerable, showing the lowest scores in quality of life and wellbeing, alongside higher stress and depression compared to retirees. These findings suggest that the 12-month window prior to retirement is a period of heightened psychological strain, mirroring qualitative reports of a ‘retirement cliff-edge’ where officers describe feeling in a state of limbo due to pension and administrative uncertainties (Vaportzis & Edwards, under review). This suggests that the anticipatory anxiety of the transition, characterised by a state of professional and financial limbo, may be more taxing than the exit itself. This aligns with the state of limbo previously noted regarding retirement dates and future security (Jones, Hesketh, & Noble, 2025a). Conversely, reaching full retirement appears associated with the most favourable psychological outcomes in this sample. Early leavers also faced significant psychological distress, reporting higher depression, anxiety, and stress than retirees, while perceiving also the lowest levels of organisational support. Importantly, soon-to-retire participants scored significantly lower on subjective wellbeing than early leavers. This suggests that the uncertainty characterising the final 12 months of service may be more psychologically taxing than the period following an early, albeit premature, exit from the Force.

Compared to the general population, while police officers demonstrated typical levels of quality of life, they presented a complex profile of heightened psychological distress alongside greater resilience and wellbeing. This suggests that while high individual resources facilitate wellbeing and quality of life, they do not erase the lasting psychological impact of the role. This phenomenon represents a ‘wellbeing paradox’, wherein, a successful transition is characterised not by the absence of psychological distress, but by officers’ ability to remain resilient despite the lingering legacy of their career. Ultimately, it appears that officers can maintain high levels of subjective functioning and wellbeing even while carrying a significant and lasting psychological burden from their career.

### Theoretical implications: The JD-R model

The JD-R model provided a robust framework for predicting post-service adjustment. In the hierarchical regression models, perceived organisational support and resilience emerged as the most consistent predictors of positive outcomes. These resources significantly predicted improved wellbeing and quality of life while reducing all measures of psychological distress. Specifically, lower levels of perceived organisational support were significantly associated with increased psychological distress. This quantitative link is reinforced by qualitative evidence suggesting a breach of the psychological contract at the exit point, where the relationship between the officer and the Force shifts from a relational bond to a transactional disposal (Vaportzis & Edwards, under review).

Resilience and perceived organisational support acted as critical buffers against the legacy of job demands. While chronic demands like trauma exposure and shift dissatisfaction initially predicted strain, their impact was significantly attenuated by these resources, confirming the JD-R model’s buffering hypothesis within the police exit context. Importantly, the ‘soon-to-retire’ status remained a significant predictor of poorer outcomes even when accounting for resources. This suggests that the anticipatory retirement period (likely exacerbated by pension uncertainty) acts as a significant demand that current internal resources may not fully buffer.

### Practical implications

The findings point toward specific areas where police organisations and policymakers could intervene. The higher psychological distress observed among soon-to-retire officers highlights a clear requirement for enhanced exit support. This group specifically identified a gap in career, financial, and pension-related guidance, with participants specifically citing the chaos of the McCloud remedy as a source of anxiety and a barrier to a smooth transition (Vaportzis & Edwards, under review). Specifically, the retrospective nature of the pension choice, and the resulting tax and administrative complexities, requires specialist financial counselling that goes beyond standard retirement signposting. Furthermore, early leavers emerged as a high-risk group for poor mental health; they reported feeling the least supported by the organisation and expressed a significantly greater need for welfare and mental health signposting than any other group.

With over half the sample welcoming career advice, financial guidance, and effective mental health signposting, the organisational mandate is clear. The urgency of this provision is highlighted by the finding that the desire for welfare signposting had the strongest relationship with negative outcomes, correlating significantly with higher levels of depression, anxiety, and stress. This identifies effective mental health signposting as the most critical intervention for mitigating psychological distress during the transition period. The significant correlation between these unmet needs and negative mental health outcomes suggests that proactive service provision, specifically targeting the 12-month transition period, could directly mitigate distress and bolster post-service wellbeing.

### Strengths, limitations, and future research

A strength of this study is the use of multiple validated scales providing high internal consistency and reliability. The collaboration with a lived-experience expert ensures the research is grounded in the practical realities of policing. In addition, the study captured a range of experiences by including retirees, early leavers, and those still serving. However, the ‘early leaver’ and ‘soon-to-retire’ groups were relatively small, therefore, comparisons should be treated as exploratory. Recruitment via social media and police-related organisations may have attracted individuals with particularly strong feelings about their transition. Finally, the study is cross-sectional, making it difficult to establish definitive causal links between service demands and long-term outcomes.

Future studies should employ longitudinal designs to track officers from their final year of service through the first several years of retirement. This would better identify the exact points at which wellbeing declines and where interventions are most effective. Additionally, investigating the specific administrative impact of the McCloud remedy on mental health would provide timely data for refining pension communications and support.

## Conclusion

This study demonstrates that the transition out of UK policing is a multidimensional process significantly impacted by the timing of exit and the presence of organisational resources. While officers exhibit remarkable resilience, the ‘wellbeing paradox’ (i.e., high subjective wellbeing alongside elevated psychological distress) highlights the lasting psychological footprint of a policing career. The 12-month window prior to retirement and the experience of leaving the Force early represent periods of acute vulnerability that current organisational resources do not fully buffer. By proactively addressing unmet needs for financial, career, and mental health support during these critical windows, police organisations can better fulfil their mandate to protect those who have served.

## Data Availability

All data produced in the present study are available upon reasonable request to the authors

## Notes

### Competing Interest Statement

The authors have declared no competing interest.

### Funding Statement

This study did not receive any funding

### Author Declarations

Ethics approval was granted by the Humanities, Social, and Health Sciences Research Ethics Panel at the University of Bradford (Reference Number E1350). All participants provided written informed consent.

